# Predicting Genetic Variant Pathogenicity Using Vector Embeddings

**DOI:** 10.1101/2025.08.08.25333318

**Authors:** Jiawei Wu, Michael Muriello, Donald G. Basel, Xiaowu Gai

**Affiliations:** Department of Pediatrics, Medical College of Wisconsin, Milwaukee, WI 53226, USA; Linda T. and John A. Mellowes Center for Genomic Sciences and Precision Medicine, Medical College of Wisconsin, Milwaukee, WI 53226, USA

**Author notes:** Correspondence to: Xiaowu Gai, PhD Department of Pediatrics, Medical College of Wisconsin Milwaukee, WI 53226.

**Keywords:** : Pathogenicity Prediction · Vector Embedding · Large Language Model · *BRCA* · *FBN1*

## Abstract

**Background:** Interpreting the pathogenicity of genetic variants remains a critical bottleneck in genomic medicine. Millions of variants of uncertain significance (VUS) hinder the clinical application of genetic findings. Traditional computational approaches often rely on hand-engineered features and fail to fully capture the complexity of multidimensional genomic annotations.

**Methods:** We developed a novel semantic embedding framework, VUS.Life, that transforms variant annotations into natural language descriptions and leverages pre-trained language models to capture pathogenicity-relevant relationships in high-dimensional vector space. This approach enables direct pathogenicity prediction through representation learning rather than traditional feature-based methods.

**Results:** We evaluated the framework using curated variants from three disease-associated genes: *BRCA1* (n=3,311) and *BRCA2* (n=4,074) from BRCA Exchange, and *FBN1* (n=1,532) from ClinVar. Variant annotations from the Variant Effect Predictor (VEP) were converted into natural language while excluding fields linked to known pathogenicity assertions to prevent data leakage. We then embedded these descriptions using three models: MPNet (all-mpnet-base-v2), Google’s text-embedding-004, and MedEmbed-large-v0.1. A k-nearest neighbor (k-NN) approach (up to 20 neighbors) was used to predict pathogenicity for new or unreviewed variants. Dimensionality reduction techniques (PCA, t-SNE, UMAP) enabled visualization of the embedding spaces.

k-NN classification showed exceptional performance across all genes and embedding models. For *BRCA1*, overall accuracy ranged from 97.3% (Google) to 97.9% (MPNet), with benign/likely benign accuracy of 95.1–97.2% and pathogenic/likely pathogenic accuracy of 97.9–98.4%. For *BRCA2*, overall accuracy ranged from 97.9% (Google) to 99.1% (MedEmbed), with benign/likely benign accuracy of 96.6–99.2% and pathogenic/likely pathogenic accuracy of 98.6–99.4%. *FBN1* validation confirmed the method’s generalizability, with accuracy exceeding 96% across all embeddings. Application to not-yet-reviewed *BRCA1/2* variants demonstrated the framework’s practicality and scalability, with unknown variants aligning closely to known benign or pathogenic clusters.

**Conclusions:** This semantic embedding framework, VUS.Life, accurately captures pathogenicity-relevant features from complex variant annotations, enabling high-accuracy (>96%) automated classification across multiple genes and models. The approach generalizes beyond well-curated genes and supports scalable, interpretable, and representation-based classification of VUS. It holds significant promise for alleviating the variant interpretation bottleneck in clinical genomics.

## 1. Introduction

### 1.1. Background

In just two decades, genomic medicine has transformed from a futuristic vision into clinical reality. What once required years and millions of dollars—sequencing a human genome—can now be accomplished in hours for under a thousand dollars [1]. This revolution has enabled genetic testing to guide cancer treatment decisions, predict disease risk, and enable precision therapies. Yet paradoxically, as our capacity to identify genetic variants has exploded, our ability to interpret their clinical significance has become the rate-limiting challenge in realizing genomic medicine’s potential.

The magnitude of this challenge is staggering. Current databases contain hundreds of millions human genetic variants, yet only a fraction carry established clinical classifications [2–4]. For genes like *BRCA1* and *BRCA2*, where accurate interpretation can mean the difference between prophylactic surgery and routine screening, this uncertainty carries profound implications.

Numerous computational tools like SIFT [5], PolyPhen-2 [6], and CADD [7], often referred to as *in silico* predictors, have been developed to help classify the pathogenicity of genetic variants by leveraging evolutionary conservation, protein structural predictions, and statistical modeling. **Table 1** lists some of the most popular and commonly used general-purpose predictors. These tools are widely used as a form of evidence in variant interpretation and are supported by clinical guidelines from the American College of Medical Genetics and Genomics (ACMG) and the Association for Molecular Pathology (AMP) [8]. Beyond these general-purpose predictors, significant research efforts have been invested in addressing challenges specific to a variant type or disease. For example, VEST-indel, focusing on insertions and deletions (indels), achieved sufficient accuracy to aid in clinical classification [9, 10]. PathoPredictor, on the other hand, is a disease-specific ensemble classifier trained on data from distinct patient cohorts, demonstrating the value of tailored models over general-purpose predictors [11].

**Table 1:**
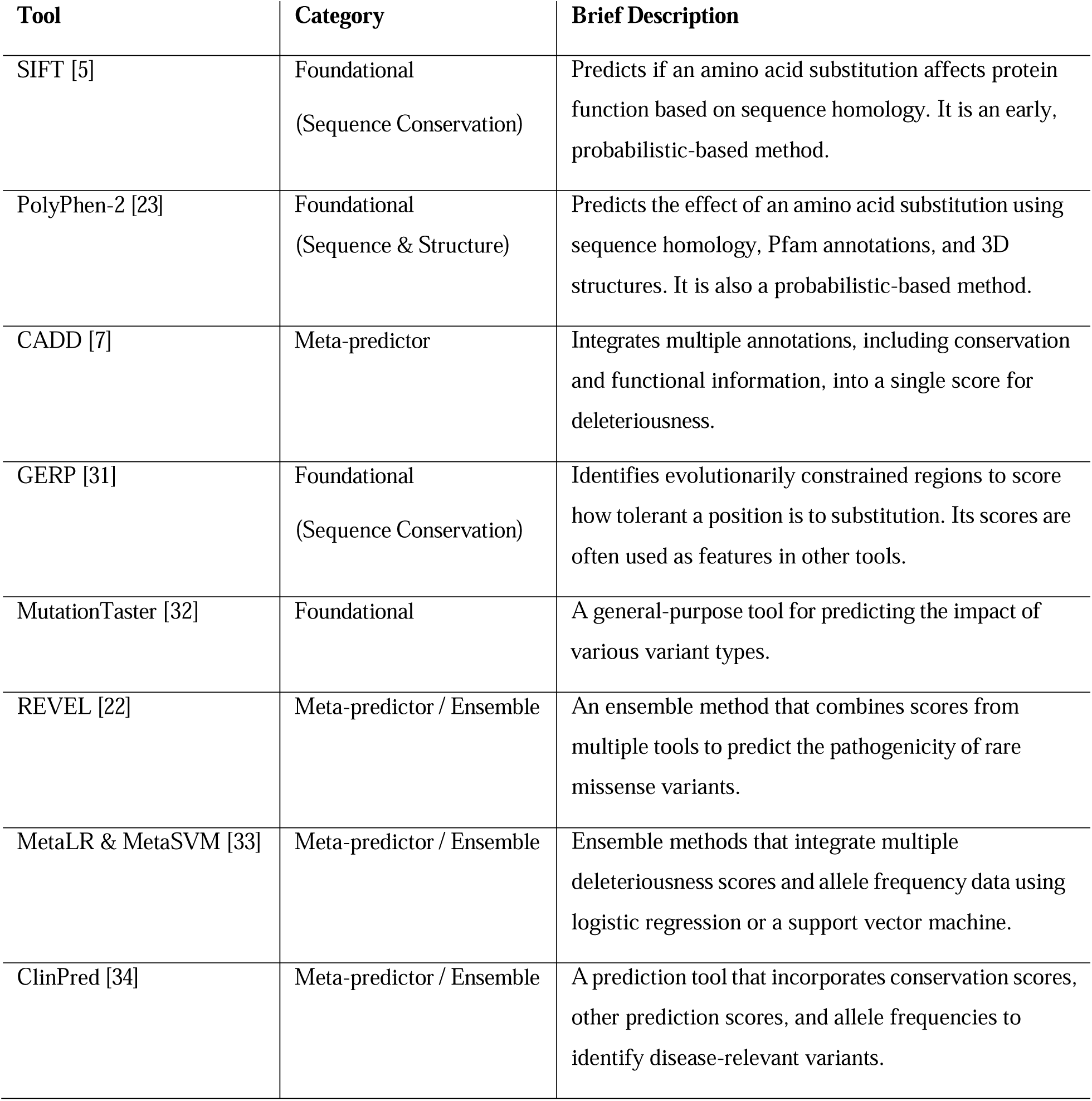
The summary of conventional methods of pathogenicity prediction.

PathoPredictor is also one of many recent predictors that have leveraged machine learning (ML) and deep learning (DL) to capture complex genomic patterns and improve predictive power. Another prominent example of the group is AlphaMissense, which adapts the state-of-the-art AlphaFold protein structure model to predict the functional impact of missense variants [12]. GNN-MAP, on the other hand, uses graph neural networks (GNNs) to model the relationships between different variants [13]. This approach enables GNN-MAP to predict pathogenicity for multiple variant types (missense, stop-gain, frameshift) and has shown strong performance on rare variants.

However, these afore-mentioned methods face inherent limitations: they typically focus on individual types of variants or features, require extensive domain expertise to engineer appropriate features, and struggle to integrate the increasingly complex landscape of genomic annotations into unified predictions.

In the last few years, Large Language Models (LLMs) have demonstrated unprecedented ability to capture semantic relationships within textual data, leading to breakthroughs in machine translation, question answering, and document understanding [14]. They have also been successfully applied in the field of genomic medicine. One application is to apply language models to the clinical narratives associated with variants. For instance, ClinVar-BERT is a model fine-tuned on millions of free-text clinical summaries from the ClinVar database [15]. By training a biomedical-specific BERT model to discern patterns in the evidence described in these reports, they successfully re-classified VUS in a way that correlated with experimental functional data. This work demonstrated that unstructured clinical text contains rich, learnable signals of pathogenicity that can be captured by language models. Another powerful approach treats biological sequences themselves as a language. Protein language models (pLMs) like ESM-1b are pre-trained on hundreds of millions of protein sequences and learn the fundamental “grammar” of protein structure and function [16]. Building on this, Lin et al. developed VariPred, which uses embeddings from a pLM to predict the impact of missense mutations [17]. By comparing the embeddings of the wild-type and mutant protein sequences, VariPred achieves state-of-the-art performance using only sequence information as input, effectively bypassing the need for explicit structural or evolutionary features. Similarly, Brandes et al. showed that the ESM1b model could be used directly, without fine-tuning, to predict the effects of all possible missense variants, in-frame indels, and even isoform-specific effects, showcasing the remarkable generalizability of these models [16].

### 1.2. A New Paradigm: Semantic Embeddings for Variant Interpretation

This study introduces a fundamentally different approach using LLMs—one that treats comprehensive genomic annotations as a structured narrative rather than discrete features. We name the framework VUS.Life. Our framework transforms variant annotations into natural language descriptions, then leverages pre-trained language models to generate semantic embeddings that capture nuanced relationships between different types of genomic evidence.

With VUS.Life, we imagine each genetic variant existing within a vast semantic space, where variants with similar functional consequences and clinical implications naturally cluster together. In this space, the pathogenicity of a newly discovered variant can be inferred from its semantic similarity to variants with established clinical significance, revealing how expert clinical geneticists integrate multiple lines of evidence and recognize patterns from experience. And, unlike most if not all aforementioned tools that focus on predicting deleteriousness of a variant, which is not equivalent to pathogenicity, VUS.Life is a framework for pathogenicity prediction directly.

## 2. Methodology

### 2.1. Development and Validation

#### Variant Datasets

To develop and validate the VUS.Life framework, we analyzed 8,917 variants with established pathogenicity classifications in hereditary breast and ovarian cancer genes *BRCA1*, *BRCA2*, and Marfan syndrome gene *FBN1*, sourced from BRCA Exchange [18] and ClinVar [19] (**Table 2**). BRCA Exchange provided established pathogenicity assertions for many highly curated variants in *BRCA1* (n = 3,311) (**Supplementary Table 1)** and *BRCA2* (n = 4,074) (**Supplementary Table 2**) genes. For *FBN1* gene, 1,532 variants were sourced from ClinVar, filtered for pathogenic, likely pathogenic, benign, and likely benign classifications, as determined by expert panel review or consensus from multiple submitters (**Supplementary Table 5**).

**Table 2:**
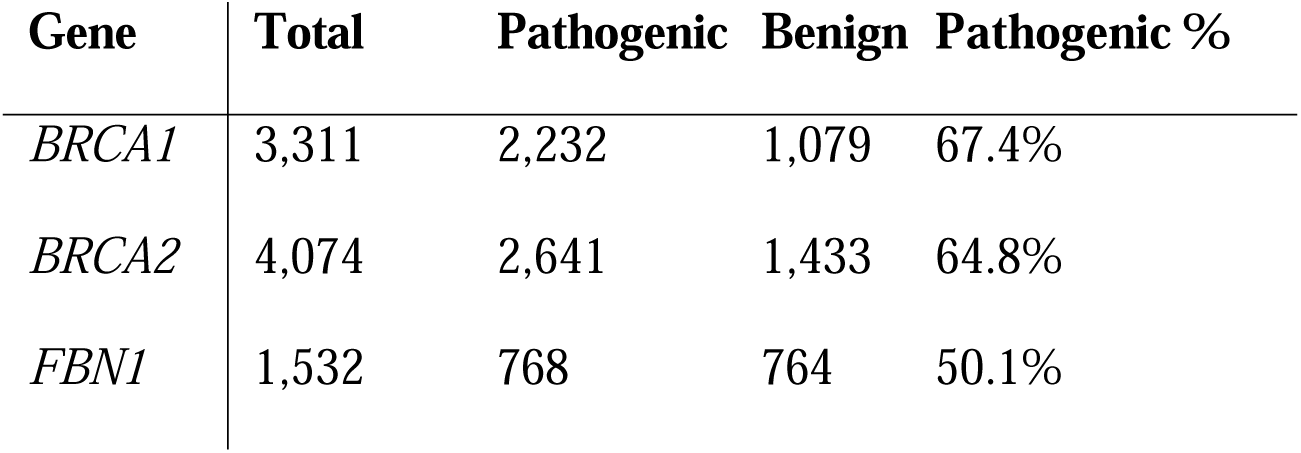
Summary of variant datasets used for model training and testing.

#### Annotation Fetch and Processing

Variants were annotated using the Variant Effect Predictor (VEP) with an extensive suite of annotation sources and prediction tools [20]. The VEP configuration was designed to capture a wide range of functional, evolutionary, and predictive information relevant to variant pathogenicity. Key annotation categories included:

- **Core Annotations**: Canonical transcript identification, predicted consequence (e.g., missense, frameshift), protein-level effects, and splicing predictions (SpliceAI [21]).
- **Pathogenicity Prediction Tools**: Scores from established predictors such as CADD [7], REVEL [22], AlphaMissense [12], SIFT [5], and PolyPhen-2 [23].
- **Regulatory and Functional Annotations**: Gene Ontology (GO) terms, pathway information, protein domain annotations, tissue-specific expression data, and loss-of-function (LoF) predictions.

To prepare the data for language model processing, we systematically converted the structured VEP JSON annotations into a consistent, semi-structured textual format. This was achieved using a custom template that serializes the nested JSON data into a human-readable, key-value report. The template organizes information into logical sections, such as variant identifiers, transcript-level consequences, population frequencies, and scores from various prediction tools (e.g., CADD, SpliceAI, AlphaMissense). This ensures uniform representation of all annotation fields. By converting numerical scores, flags, and identifiers into a unified textual string, we create a comprehensive and uniform input format that enables the embedding model to learn the semantic relationships between disparate types of genomic evidence. An abbreviated example of the generated text for an intronic variant is shown below: *Variant ID: NC_000017.11:g.43046253G>A. Variant class: SNV. Most severe consequence: intron_variant. Transcript consequence #1: The transcript id is NM_007294.4; The gene symbol is BRCA1; The gene id is 672; The gene symbol source is EntrezGene; The biotype is protein_coding. SpliceAI scores: The SpliceAI SYMBOL is BRCA1; The SpliceAI DS AG is 0.0; The SpliceAI DS AL is 0.0. Population frequencies: Allele: A. The frequency in gnomadg_asj is 0.02244; The frequency in amr is 0.0043; The frequency in gnomadg_ami is 0.008216; The frequency in gnomadg_afr is 0.002173*.

### 2.2. Semantic Embedding Models and Generation

Three distinct embedding models were selected to capture different aspects of semantic representation:

- **all-mpnet-base-v2** (MPNet) [24]: A Transformer-based model pre-trained using masked and permuted language modeling. With 768 dimensions and a 512-token limit, it excels at semantic similarity tasks, balancing sentence structure understanding.
- **MedEmbed-large-v0.1** (MedEmbed) [25]: A domain-specific Transformer trained on biomedical literature, offering 1,024 dimensions and a 512-token limit. Its specialization provides a nuanced understanding of medical and genomic terminology.
- **text-embedding-004** (Google Embeddings) [26]: Google’s state-of-the-art general-purpose embedding model, generating 768-dimensional embeddings with a 2,048-token limit, performing exceptionally well on diverse semantic tasks.

The embedding generation process transforms each variant’s description into vector representations using these models. These embeddings are stored in <u>ChromaDB (</u>https://github.com/chroma-core/chroma), a vector database optimized for similarity search and retrieval. Accompanying metadata includes variant identifiers, genomic coordinates, pathogenicity classifications, confidence scores, gene annotations, functional consequences, and processing timestamps. Vector indices are tuned for cosine similarity search, with parameters adjusted to each model’s dimensionality.

### 2.3. Evaluation

#### 2.3.1. Embedding Space Visualization and Analysis

To analyze the high-dimensional embeddings, we applied three complementary dimensionality reduction techniques: Principal Component Analysis (PCA) [27], t-Distributed Stochastic Neighbor Embedding (t-SNE) [28], and Uniform Manifold Approximation and Projection (UMAP) [29]. PCA projects data into a visualizable space by maximizing variance, t-SNE emphasizes local neighborhood relationships, and UMAP balances local and global structure preservation. These visualizations reveal distinct clusters of variants with shared pathogenicity, indicating that the embedding models effectively capture pathogenicity-related features. The discriminative power was quantified by correlating embedding distances with pathogenicity differences, confirming tighter clustering of variants with similar clinical impacts. This structure also supports the k-NN classification approach for predicting pathogenicity of novel variants or variants of uncertain significance.

#### 2.3.2. Quantification of Model Performance

A k-NN classification approach was developed to predict variant pathogenicity using semantic embeddings. Performance was evaluated across multiple neighborhood sizes (k = 1, 5, 10, 15, 20) to identify gene-specific optimal k values. Cosine similarity was employed as the distance metric due to its effectiveness in high-dimensional spaces and its robustness to vector magnitude variations. For each query variant, the algorithm identifies the k most similar variants with known pathogenicity, excluding the query variant from the training set to prevent data leakage. Predictions are based on majority voting, with confidence scores reflecting the proportion of votes supporting the predicted class. The embedding space’s ability to separate variants by pathogenicity was assessed by computing a clustering accuracy metric for known variants. For each variant with established pathogenicity, its k nearest neighbors were identified, and the percentage of neighbors sharing the same classification was calculated. The classification criteria are outlined in **Table 3**.

**Table 3:** Criteria for classifying pathogenicity based on the proportion of pathogenic and benign neighbors among the top-k nearest neighbors.

| **Classification** | **Criteria (among k neighbors)** | **Confidence Score** |
| --- | --- | --- |
| **Potentially Pathogenic** | $\geq 90\%$ pathogenic neighbors | Pathogenic Ratio |
| **Likely Pathogenic** | $\geq 60\%$ pathogenic neighbors (but $< 90\%$) | Pathogenic Ratio |
| **Potentially Benign** | $\geq 90\%$ benign neighbors | Benign Ratio |
| **Likely Benign** | $\geq 60\%$ benign neighbors (but $< 90\%$) | Benign Ratio |
| **Uncertain** | Mixed or no clear majority | Max (benign, pathogenic) Ratio |

**Table 4:** Clustering accuracy of semantic embedding models for predicting variant pathogenicity across genes. Accuracy is measured by the percentage of k-nearest neighbors sharing the same known pathogenicity classification. Higher accuracy indicates better separation of pathogenic and benign variants.

| Gene | Model | k=5 | k=10 | k=15 | k=20 |
| --- | --- | --- | --- | --- | --- |
| *BRCA1* | MedEmbed-large-v0.1 | 0.981 | 0.980 | 0.978 | 0.977 |
|  | google-embedding | 0.980 | 0.977 | 0.974 | 0.973 |
|  | all-mpnet-base-v2 | 0.984 | 0.983 | 0.981 | 0.979 |
| *BRCA2* | MedEmbed-large-v0.1 | 0.993 | 0.993 | 0.992 | 0.991 |
|  | google-embedding | 0.985 | 0.983 | 0.981 | 0.979 |
|  | all-mpnet-base-v2 | 0.992 | 0.991 | 0.990 | 0.990 |
| *FBN1* | MedEmbed-large-v0.1 | 0.966 | 0.965 | 0.964 | 0.963 |
|  | google-embedding | 0.965 | 0.964 | 0.961 | 0.959 |
|  | all-mpnet-base-v2 | 0.967 | 0.962 | 0.961 | 0.959 |

### 2.4. Implementation

The pathogenicity prediction pipeline targets novel and variants of uncertain significance (VUS) using a modular, efficient workflow. Unknown variants are retrieved from ChromaDB (https://github.com/chroma-core/chroma) using pathogenicity status filters, processed in batches for computational efficiency, and analyzed for their k (default: 20) nearest known variants (benign or pathogenic) via cosine similarity. Predictions are generated using a confidence-weighted voting system, with confidence scores reflecting prediction reliability. Visualizations employing PCA, UMAP, and t-SNE illustrate spatial relationships between known and unknown variants, aiding genetic counselors and clinicians in interpreting results.

## 3. Results

### 3.1. Embedding Results

The embedding analysis establishes a robust framework for clinical variant interpretation via confidence-based stratification derived from neighborhood consistency patterns. Variants can be categorized as high-confidence (>95% agreement among nearest neighbors), moderate-confidence (60-95% agreement), or boundary cases requiring expert review (<60% agreement). *BRCA1* analysis revealed that 2-5% of variants fell into boundary cases across models, highlighting cases that may benefit from additional clinical review or functional studies. *BRCA2* showed fewer boundary cases (0.4-2.6%), indicating stronger semantic clustering and higher prediction confidence. The gradient of neighborhood agreement serves as a natural quality control mechanism for clinical workflows, enabling automated processing of high-confidence predictions while flagging uncertain cases for genetic counseling and expert interpretation. This approach addresses the critical need for reliable variant classification in hereditary cancer predisposition testing, where accurate interpretation directly influences clinical management decisions. Consistent performance across neighbor counts (k=1, 5, 20) validates the methodology’s robustness, with optimal k likely being gene-specific to balance local precision and noise reduction for immediate clinical implementation.

#### *BRCA1* Semantic Embedding Performance

The k-nearest neighbors (K-NN) evaluation of *BRCA1* variants (n=3,311) demonstrated robust classification performance across all three embedding models, with systematic analysis confirming the effectiveness of semantic representations in capturing pathogenicity-relevant features (**Figure 1**). The MPNet model achieved the highest overall accuracy at 97.9% with top 20 neighbors, with strong performance for both benign/likely benign variants (96.9%) and pathogenic/likely pathogenic variants (98.4%). MedEmbed showed comparable performance at 97.7% overall accuracy, while Google embeddings achieved 97.3% overall accuracy. Neighborhood consistency analysis revealed that 94.6% of benign variants and 97.4% of pathogenic variants had 80-100% agreement among their top 20 nearest neighbors when using MPNet, indicating robust semantic clustering. The high neighborhood consensus across all models suggests that *BRCA1* variant annotations contain sufficient semantic information to enable reliable automated classification. MedEmbed exhibited 92.5% consistency for benign variants achieving 80-100% neighborhood agreement, while Google embeddings achieved 92.6% consistency for benign variants. This consistent performance across different embedding approaches validates the robustness of the semantic representation framework for *BRCA1* variant interpretation.

**Figure 1:**
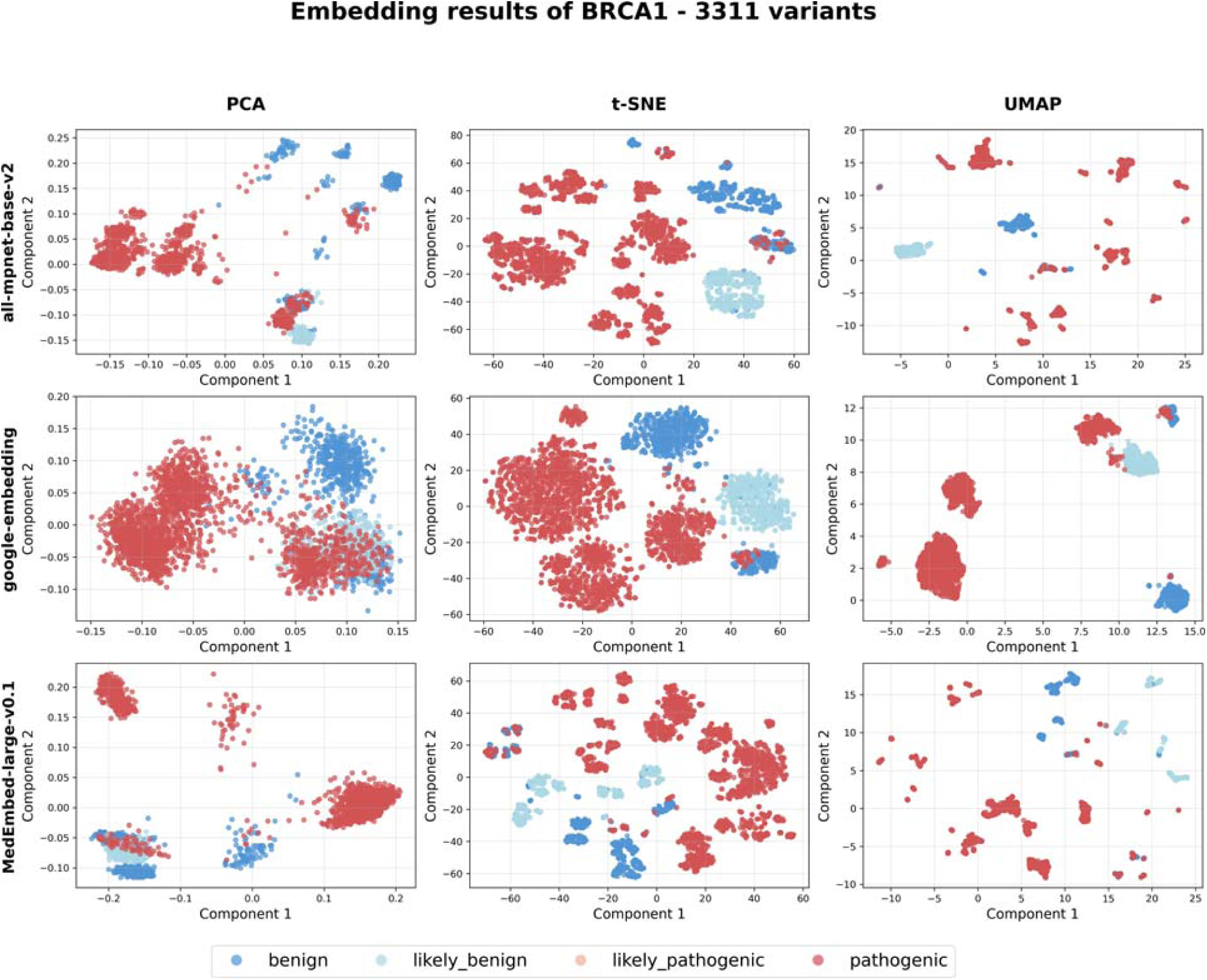
Dimensionality reduction of semantic embeddings for 3,311 *BRCA1* variants. Each panel displays a 2D projection of the high-dimensional variant embeddings using three different models (rows: all-mpnet-base-v2, google-embedding, MedEmbed-large-v0.1) and three dimensionality reduction techniques (columns: PCA, t-SNE, UMAP). The distinct separation between benign (blue) and pathogenic (red) variants across all models demonstrates that the embeddings effectively capture pathogenicity-relevant features. Non-linear methods like t-SNE and UMAP reveal more defined clusters compared to the linear PCA projection, confirming the semantic space is structured by clinical significance.

#### *BRCA2* Semantic Embedding Performance

Our method demonstrated exceptional classification performance on *BRCA2* variants (n=4,074) across all embedding models, with MedEmbed achieving outstanding results of 99.1% overall accuracy (99.2% benign/likely benign, 99.1% pathogenic/likely pathogenic) (**Figure 2**). The domain-specific medical embedding model’s superior performance highlights the value of specialized pre-training for genetic variant analysis. MPNet achieved comparable excellence with 99.0% overall accuracy (98.3% benign/likely benign, 99.4% pathogenic/likely pathogenic), while Google embeddings maintained strong performance at 97.9% overall accuracy. Neighborhood distribution analysis revealed remarkable consistency, with MedEmbed showing 99.6% of benign variants and 98.8% of pathogenic variants achieving 80-100% agreement among top 20 neighbors. MPNet demonstrated equally impressive consistency with 97.4% of benign variants and 99.3% of pathogenic variants showing high neighborhood agreement. The larger sample size for *BRCA2* variants appears to contribute to enhanced model training and more stable semantic representations, enabling the development of highly reliable prediction models for clinical applications. Google embeddings, while achieving the lowest performance among the three models, still maintained clinically relevant accuracy with 94.9% of benign variants showing strong neighborhood consensus.

**Figure 2:**
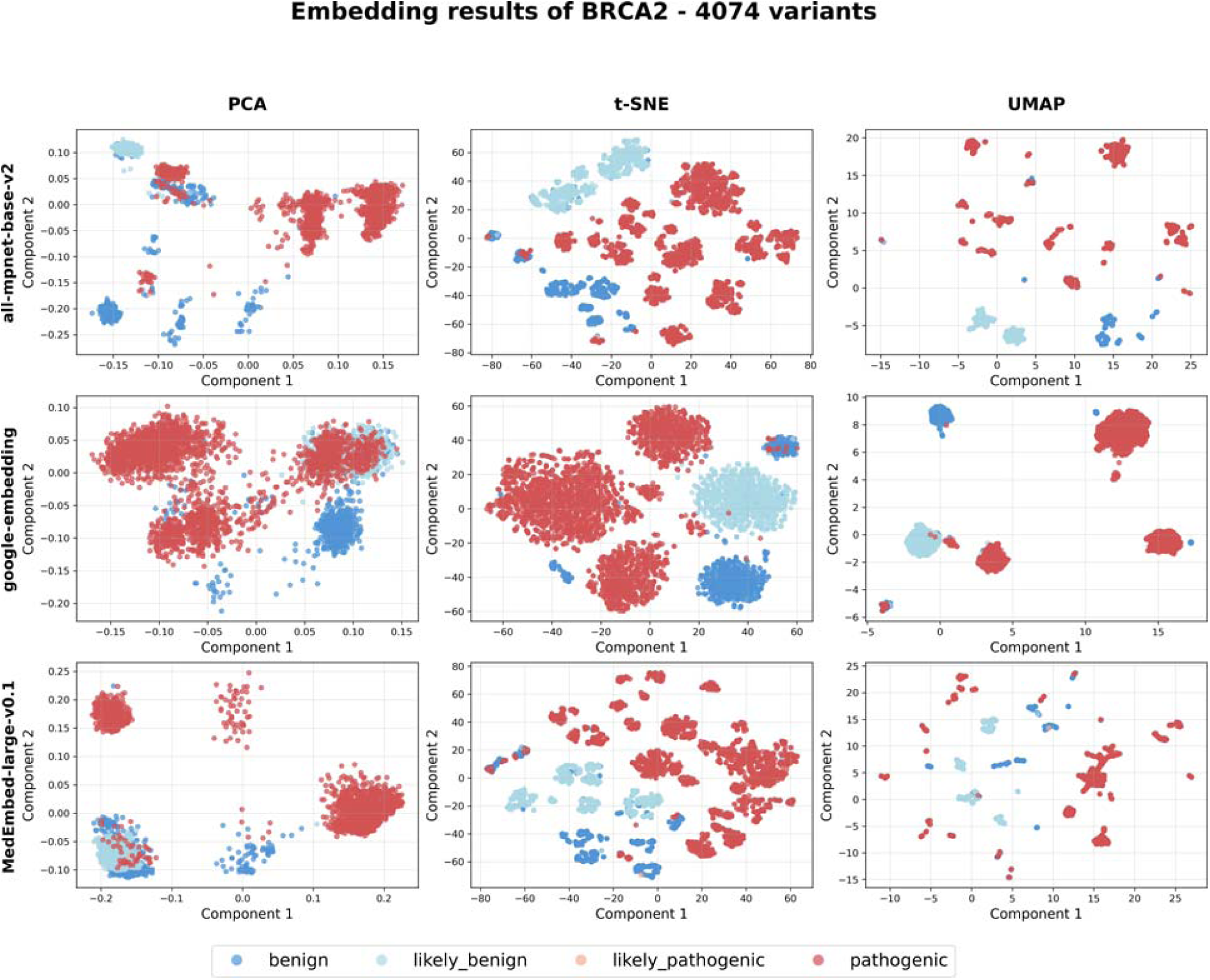
Visualization of semantic embeddings for 4,074 *BRCA2* variants. Following the same structure as Figure 1, these plots show the embedding space for *BRCA2* variants. The separation between benign (blue) and pathogenic (red) classes is even more pronounced than for *BRCA1*, with t-SNE and UMAP revealing highly compact and well-separated clusters, suggesting a very robust semantic representation.

#### *FBN1* Semantic Embedding Performance

The *FBN1* validation results demonstrate robust performance across all three embedding models, achieving consistently high accuracy rates exceeding 96% (MPNet: 96.7%, Google: 96.5%, MedEmbed: 96.6% at k=5) (**Figure 3**). These results are particularly significant as they validate the generalizability of our semantic embedding approach beyond the extensively characterized *BRCA* cancer predisposition genes to a gene associated with connective tissue disorders and mostly missense instead of loss-of-function pathogenic variants. The dimensional reduction visualizations reveal distinct clustering patterns for *FBN1* variants, with Google embeddings showing the most compact and well-separated clusters in t-SNE and UMAP projections, while MPNet and MedEmbed demonstrated more distributed but still discriminative patterns. Notably, the *FBN1* performance closely mirrors that observed for *BRCA1* (97.3% with MPNet), suggesting that the embedding approach maintains effectiveness across different genetic contexts and disease mechanisms, even for genes that may have less comprehensive functional annotation compared to the intensively studied *BRCA1/2* genes. This validation supports the broader clinical utility of semantic embeddings for variant interpretation across diverse genetic conditions, particularly important for rare disease contexts where traditional computational approaches may be limited by sparse training data.

**Figure 3:**
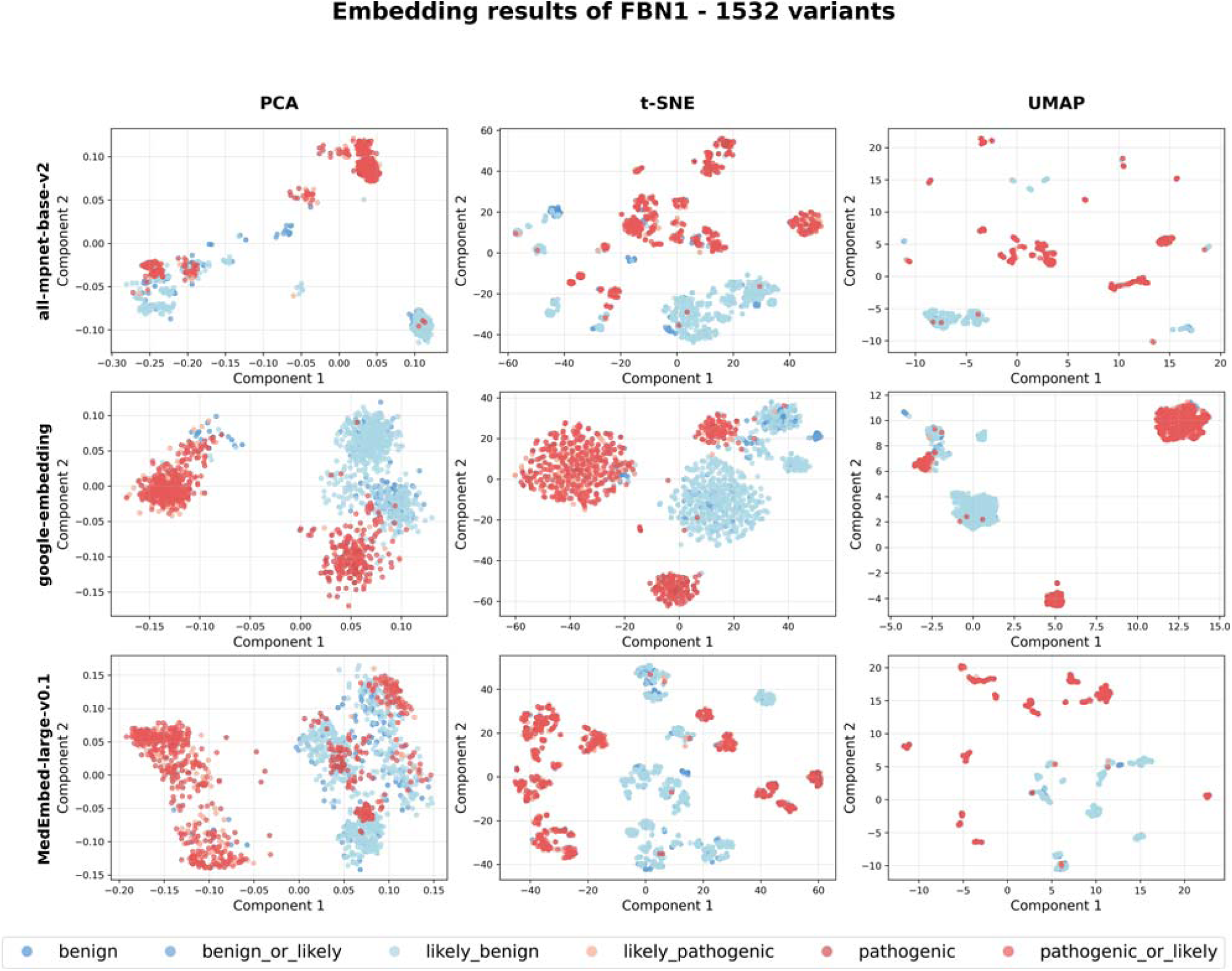
Validation of the semantic embedding framework’s generalizability using 1,532 *FBN1* variants. This figure demonstrates that the methodology is not limited to *BRCA* genes. Despite the different genetic context and disease mechanism (Marfan syndrome), the embeddings for *FBN1* variants maintain a strong separation between benign (blue) and pathogenic (red) classes. This result validates the robustness and broad utility of using semantic embeddings for variant interpretation across diverse, clinically important genes.

### 3.2. Pathogenicity Prediction

The prediction results for 3000 not-yet-reviewed variants in both *BRCA1* and 3,000 not-yet-reviewed variants *BRCA2* demonstrate the promising potential of semantic embeddings for variant pathogenicity classification (**Supplementary Tables 3 & 4**) (see also **Figures 4 & 5**). Across all three embedding models (MPNet, Google embeddings, and MedEmbed), the unknown variants (green diamonds) exhibit spatial distributions that closely mirror the established clustering patterns of known benign (blue) and pathogenic (red) variants.

**Figure 4:**
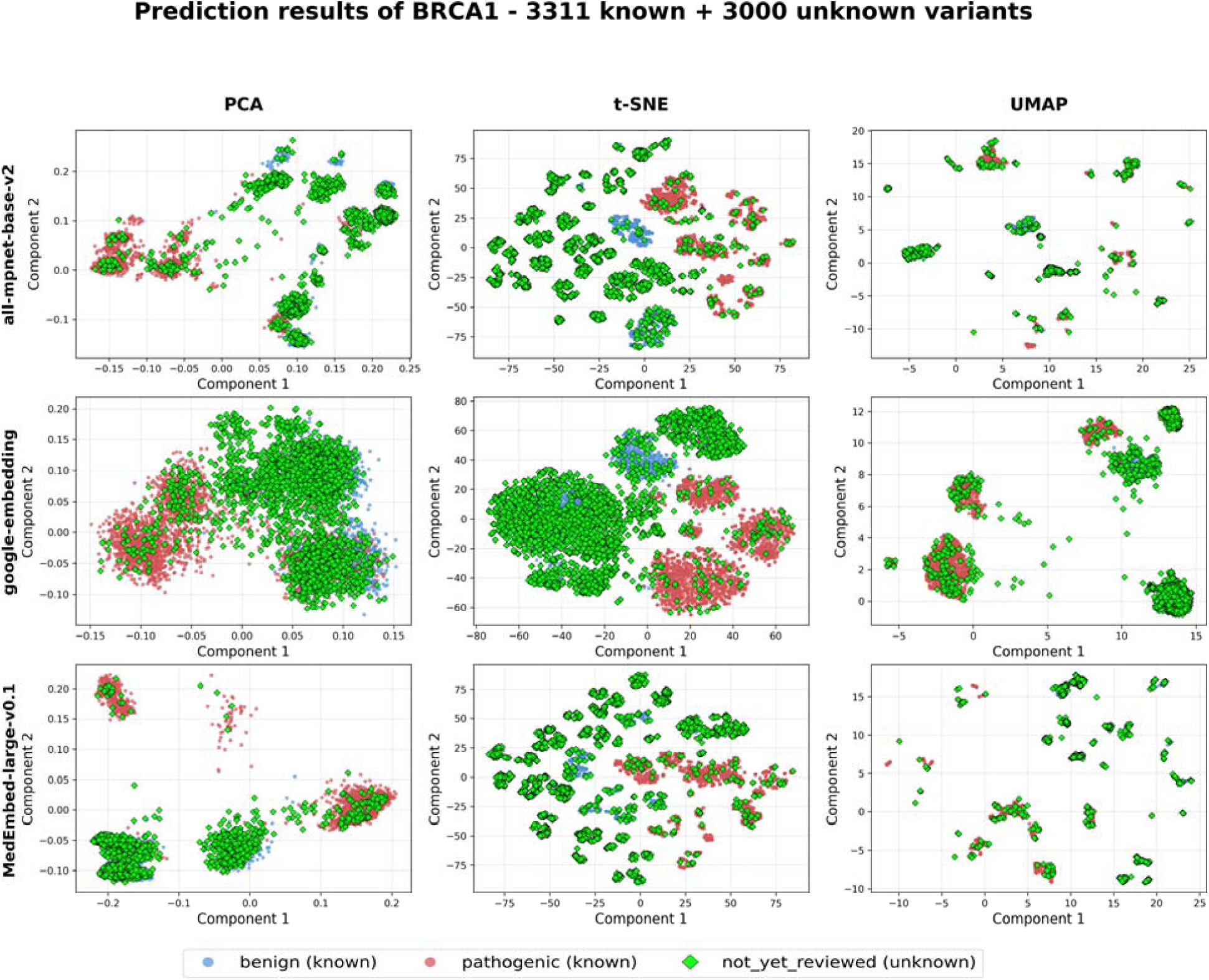
Visualization of pathogenicity of 3,000 not-yet-reviewed variants (highlighted as green diamonds) on *BRCA1*. The pathogenicity is simply mapped into binary labels: benign and pathogenic. Each panel displays dimensionality reduction (PCA, UMAP, and t-SNE) of variant embeddings. The consistent spatial distribution of unknown variants relative to known benign and pathogenic variants across models and genes suggests that embeddings effectively capture pathogenicity-related semantics.

**Figure 5:**
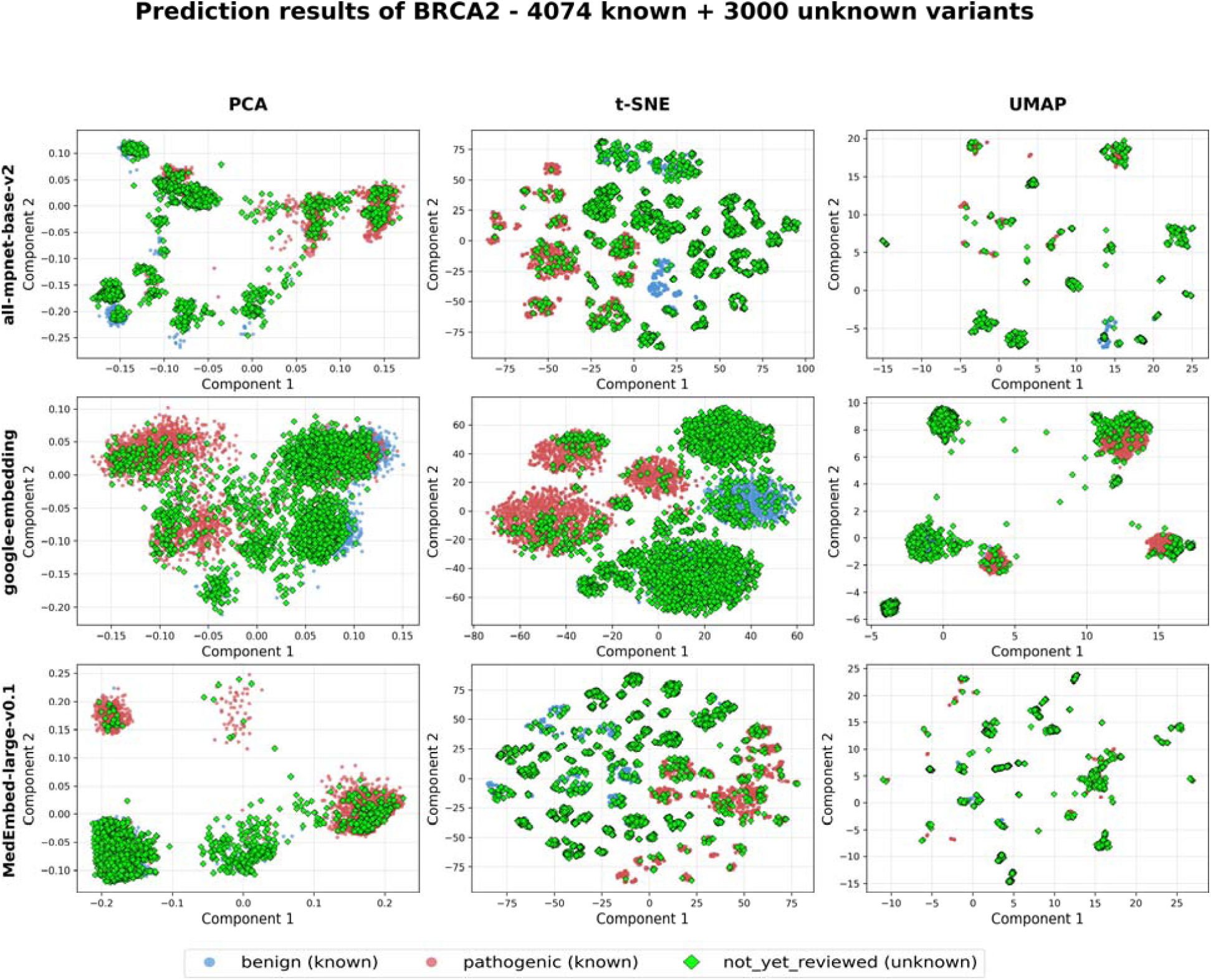
Visualization of pathogenicity of 3,000 not-yet-reviewed *BRCA2* variants. The same pattern as *BRCA1* prediction.

To illustrate the practical application of this approach, we examined prediction results from 10 randomly selected variants each from *BRCA1* and *BRCA2* datasets (**Tables 5 & 6**). The results demonstrate strong model concordance and biologically coherent predictions across different variant types. For *BRCA1* variants, all three embedding models achieved perfect agreement on clearly deleterious variants such as frameshift (g.32396971_32396972dup) and stop-gained variants (g.32339409C>G), both receiving unanimous pathogenic predictions with 1.00 confidence scores based on k-nearest neighbor counts of B:0, P:20. Intronic variants consistently received benign classifications across all models with maximum confidence, while missense variants showed more nuanced predictions with generally high confidence but occasional inter-model variation in neighbor compositions.

**Table 5:** Predicted pathogenicity of randomly selected unknown variants in *BRCA1* (NC_000017.11) using different semantic embedding models.

| Genomic HGVS | Consequence | MedEmbed |  |  | google |  |  | MPNet |  |  |
| --- | --- | --- | --- | --- | --- | --- | --- | --- | --- | --- |
|  |  | Count | Conf. Score | Pred | Count | Conf. Score | Pred | Count | Conf. Score | Pred |
| g.43094629_43095075del | splice_acceptor_variant | B:16, P:4 | 0.800 | benign | B:2, P:18 | 0.900 | pathogenic | B:10, P:10 | 0.500 | uncertain |
| g.43102853C>G | intron_variant | B:20, P:0 | 1.000 | benign | B:20, P:0 | 1.000 | benign | B:20, P:0 | 1.000 | benign |
| g.43067120C>T | intron_variant | B:20, P:0 | 1.000 | benign | B:20, P:0 | 1.000 | benign | B:20, P:0 | 1.000 | benign |
| g.43093771_43093774delinsTC | frameshift_variant | B:0, P:20 | 1.000 | pathogenic | B:0, P:20 | 1.000 | pathogenic | B:0, P:20 | 1.000 | pathogenic |
| g.43061638A>G | intron_variant | B:20, P:0 | 1.000 | benign | B:20, P:0 | 1.000 | benign | B:20, P:0 | 1.000 | benign |
| g.43065397G>A | intron_variant | B:20, P:0 | 1.000 | benign | B:19, P:1 | 0.950 | benign | B:20, P:0 | 1.000 | benign |
| g.43067430del | intron_variant | B:20, P:0 | 1.000 | benign | B:8, P:12 | 0.600 | pathogenic | B:20, P:0 | 1.000 | benign |
| g.43074502G>T | missense_variant | B:17, P:3 | 0.850 | benign | B:13, P:7 | 0.650 | benign | B:16, P:4 | 0.800 | benign |
| g.43094690T>C | missense_variant | B:20, P:0 | 1.000 | benign | B:18, P:2 | 0.900 | benign | B:20, P:0 | 1.000 | benign |
| g.43115774T>G | missense_variant | B:12, P:8 | 0.600 | benign | B:5, P:15 | 0.750 | pathogenic | B:16, P:4 | 0.800 | benign |

**Table 6:** Predicted pathogenicity of randomly selected unknown variants in *BRCA2* (NC_000013.11) using different semantic embedding models.

| Genomic HGVS | Consequence | MedEmbed |  |  | google |  |  | MPNet |  |  |
| --- | --- | --- | --- | --- | --- | --- | --- | --- | --- | --- |
|  |  | Count | Conf. Score | Pred | Count | Conf. Score | Pred | Count | Conf. Score | Pred |
| g.32353730A>G | intron_variant | B:20, P:0 | 1.000 | benign | B:20, P:0 | 1.000 | benign | B:20, P:0 | 1.000 | benign |
| g.32340247G>T | missense_variant | B:18, P:2 | 0.900 | benign | B:20, P:0 | 1.000 | benign | B:20, P:0 | 1.000 | benign |
| g.32363243A>G | missense_variant | B:19, P:1 | 0.950 | benign | B:15, P:5 | 0.750 | benign | B:20, P:0 | 1.000 | benign |
| g.32396971_32396972dup | frameshift_variant | B:0, P:20 | 1.000 | pathogenic | B:0, P:20 | 1.000 | pathogenic | B:0, P:20 | 1.000 | pathogenic |
| g.32370904A>T | intron_variant | B:20, P:0 | 1.000 | benign | B:20, P:0 | 1.000 | benign | B:20, P:0 | 1.000 | benign |
| g.32322227A>G | intron_variant | B:20, P:0 | 1.000 | benign | B:20, P:0 | 1.000 | benign | B:20, P:0 | 1.000 | benign |
| g.32385434_32385445dup | intron_variant | B:20, P:0 | 1.000 | benign | B:15, P:5 | 0.750 | benign | B:20, P:0 | 1.000 | benign |
| g.32369761A>T | intron_variant | B:20, P:0 | 1.000 | benign | B:20, P:0 | 1.000 | benign | B:20, P:0 | 1.000 | benign |
| g.32380102G>A | synonymous_variant | B:20, P:0 | 1.000 | benign | B:18, P:2 | 0.900 | benign | B:20, P:0 | 1.000 | benign |
| g.32339409C>G | stop_gained | B:0, P:20 | 1.000 | pathogenic | B:0, P:20 | 1.000 | pathogenic | B:0, P:20 | 1.000 | pathogenic |

The *BRCA2* results revealed similar patterns but with some instructive disagreements that highlighted the method’s sensitivity to semantic context. Most notably, a splice acceptor variant (g.43094629_43095075del) demonstrated model disagreement, with MedEmbed predicting benign (B:16, P:4, confidence 0.800), Google embeddings predicting pathogenic (B:2, P:18, confidence 0.900), and MPNet remaining uncertain (B:10, P:10, confidence 0.500). This variant exemplifies cases where semantic interpretation of clinical annotations may vary, potentially reflecting different emphasis on splice site disruption versus other contextual factors in the training literature.

For *BRCA1*, the unknown variants distribute throughout the embedding space in a manner consistent with the trained pathogenicity classes. In the MPNet and Google embedding visualizations, unknown variants segregate into regions predominantly occupied by either benign or pathogenic training variants, particularly evident in the t-SNE projections, where distinct clusters emerge. The MedEmbed results show a more dispersed but still meaningful distribution, with unknown variants positioning themselves in semantically appropriate regions of the embedding space.

Similar patterns are observed for *BRCA2*, where unknown variants demonstrate clear spatial alignment with their predicted pathogenicity classes. The Google embeddings show particularly strong clustering behavior, with unknown variants forming coherent groups that align with either the benign or pathogenic regions. The UMAP projections across all models reveal that unknown variants maintain the same topological relationships as the training data, suggesting robust semantic capture of pathogenicity-relevant features.

The 2015 ACMG/AMP guidelines for variant interpretation do not fully account for gene-specific characteristics. Since 2019, the ClinGen *FBN1* Variant Curation Expert Panel (VCEP), like many other VCEPs, has invested substantial effort in developing consensus recommendations tailored to the unique features of this disease gene and its pathogenic variants [30]. To evaluate these new guidelines, the panel conducted a pilot study on 60 representative yet highly challenging variants. This effort involved collaboration among three core sites and six non-core institutions, demonstrating improved interpretive consistency compared to the 2015 ACMG/AMP framework. Of these 60 variants, 30 overlapped with the 1,532 variants used to construct the *FBN1* vector embedding space. Pathogenicity prediction was therefore performed only on the remaining 30 variants. These 30 unresolved variants segregated into established pathogenic or benign clusters, particularly evident in the UMAP projection using MPNet embeddings (**Figure 6**). All 30 variants received either pathogenic or benign classifications (**Supplementary Table 6**). For 10 variants, the *FBN1* VCEP—either at core sites, non-core sites, or both—had been assigned a VUS classification under the updated guidelines. Our algorithm, using the best-performing MPNet model for *FBN1*, classified 7 of these as pathogenic and 3 as benign. For the remaining 20 variants, MPNet predictions agreed with VCEP classifications for 17 (85%). We examined the three discordant calls, which are all missense variants predicted as pathogenic by our algorithm with MPNet model but classified as benign by the FBN1 VCEP. Notably, ClinVar lists all three as VUS, with submissions from more than four independent laboratories each, underscoring the uncertainty surrounding their true pathogenicity.

**Figure 6:**
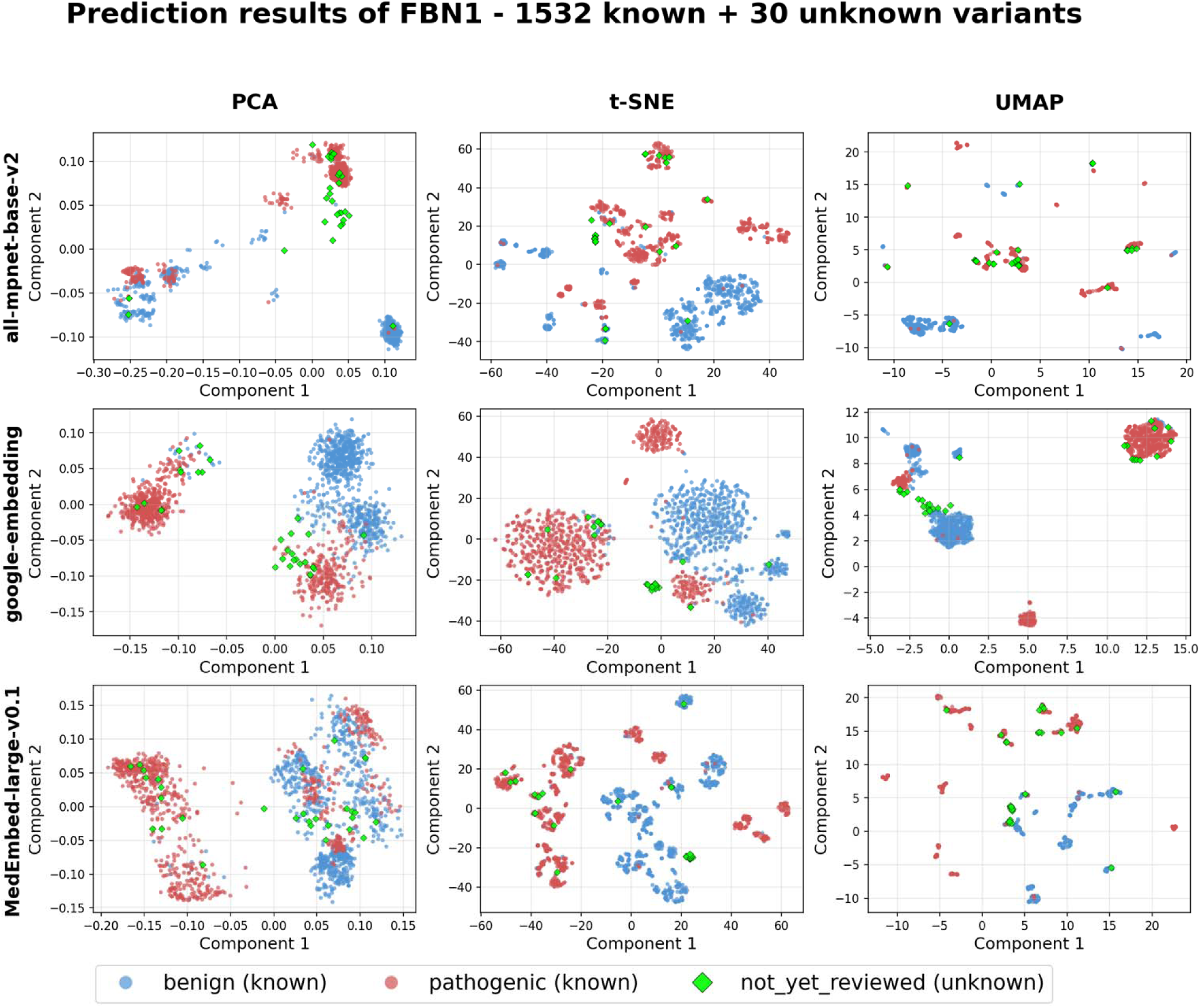
Visualization of predicted pathogenicity of 30 variants (highlighted as green diamonds) on *FBN1,* piloted by the ClinGen *FBN1* variant curation expert panel (VCEP). The pathogenicity is simply mapped into binary labels: benign and pathogenic. Each panel displays dimensionality reduction (PCA, UMAP, and t-SNE) of variant embeddings. The consistent spatial distribution of unknown variants relative to known benign and pathogenic variants across models and genes suggests that embeddings effectively capture pathogenicity-related semantics.

### 3.3. Distance Ratio Analysis Confirms Robust Class Separation

To quantitatively validate the class separation observed in the dimensionality reduction plots, we applied our Distance Ratio metric to the embeddings of *BRCA1* and *BRCA2* variants. The results confirm that all three embedding models produce a highly structured and discriminative semantic space.

As shown in **Figure 7**, the distributions of the Distance Ratio for both pathogenic (red) and benign (blue) variants are overwhelmingly concentrated to the left of the 1.0 threshold. This visually demonstrates that for most variants, the distance to same-class neighbors is significantly smaller than the distance to different-class neighbors.

**Figure 7:**
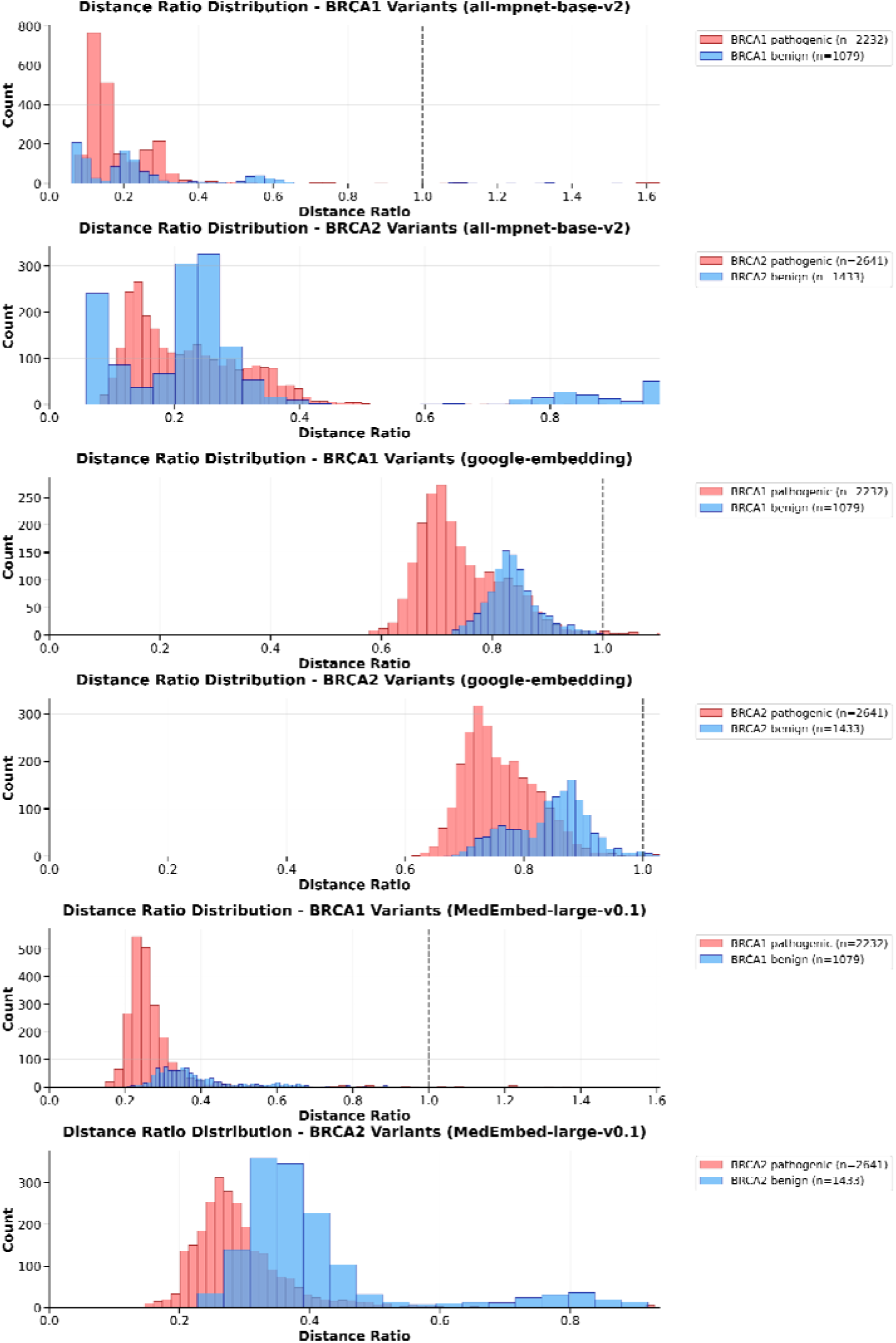
Distance Ratio distributions confirm strong separation of benign and pathogenic variants. The figure shows histograms of the Distance Ratio for *BRCA1* (top row of each panel) and *BRCA2* (bottom row) variants, evaluated on three different embedding models. The ratio compares a variant’s distance to same-class neighbors versus different-class neighbors. The dashed vertical line marks the theoretical boundary of 1.0; ratios below this line indicate effective clustering. The overwhelming concentration of both pathogenic (red) and benign (blue) distributions at ratios significantly less than 1.0 provides strong quantitative evidence that the semantic embedding space effectively separates variants based on their clinical significance.

This finding is supported by quantitative analysis (**Table 7**), which shows that across all models and genes, over 98% of variants have a Distance Ratio less than 1.0, indicating effective clustering. The all-mpnet-base-v2 model showed the most consistent high performance across all categories, with an overall average of 98.7% of variants correctly clustered. While all models performed exceptionally well, we observed that *BRCA2* variants, which have more extensive annotation data, showed slightly better class separation than *BRCA1* variants. This robust quantitative validation provides high confidence in using the semantic embedding space for downstream classification and similarity search tasks.

**Table 7:**
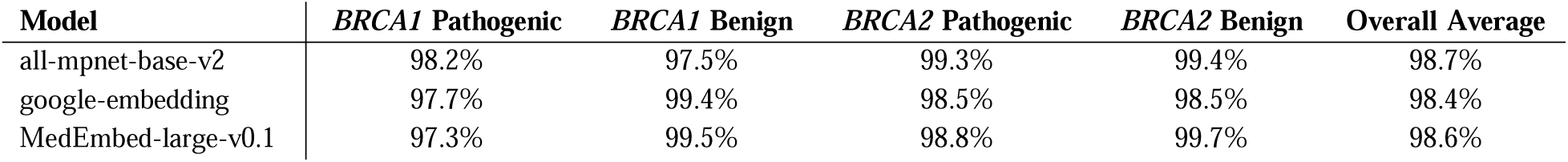
Percentage of variants with a Distance Ratio < 1.0, calculated with k=20 nearest neighbors. Values represent the proportion of variants that are closer to their same-class neighbors than to opposite-class neighbors, indicating effective separation.

#### Clinical Implications

The consistent distribution patterns across both genes and all embedding models strongly support the hypothesis that semantic embeddings can effectively capture the linguistic and clinical nuances present in variant annotations that distinguish pathogenic from benign variants. This spatial coherence between known and unknown variants indicates that the learned representations encode biologically meaningful features rather than spurious correlations, providing confidence in the method’s ability to generalize to variants of uncertain significance (VUS). The preservation of clustering structure suggests that clinicians could potentially leverage these embeddings not only for binary classification but also for confidence estimation and identification of variants requiring additional functional studies.

## 4. Discussion

This work presents a comprehensive evaluation of semantic embedding approaches for automated variant pathogenicity classification, focusing on *BRCA1* (n=3,311 variants) and *BRCA2* (n=4,074 variants) genes using three distinct embedding models: MPNet (all-mpnet-base-v2), Google embeddings, and MedEmbed (MedEmbed-large-v0.1). Our results demonstrate that semantic embeddings can effectively capture pathogenicity-relevant features from variant annotations, achieving consistently high accuracy rates exceeding 97% across all models and genes through k-nearest neighbors’ classification.

The superior clustering performance observed for *BRCA2* compared to *BRCA1* variants across all dimensionality reduction techniques (PCA, t-SNE, and UMAP) suggests that annotation quality and clinical characterization depth may significantly influence embedding effectiveness. This finding is particularly relevant given that *BRCA1* and *BRCA2* are among the most extensively studied genes in clinical genetics, with decades of functional and clinical research contributing to comprehensive variant annotations.

To validate the generalizability of our approach, we extended our analysis to *FBN1* (fibrillin-1) (n=1,532 variants), a gene associated with Marfan syndrome and related connective tissue disorders, with pathogenic variants being mostly missense instead of loss-of-function. Preliminary results indicate similarly high classification accuracy, suggesting that the semantic embedding approach maintains effectiveness across different genetic contexts and disease mechanisms. Building on this, we applied our algorithm to 30 challenging, unresolved variants from the FBN1 Variant Curation Expert Panel (VCEP) pilot study. This set included 20 variants classified as either pathogenic or benign based on the updated *FBN1* VCEP guidelines and 10 variants still remained as variants of uncertain significance (VUS). Our analysis showed that these unresolved variants consistently segregated into established pathogenic or benign clusters within the embedding spaces, enabling our method to assign pathogenicity for all 30 variants. For the 20 VCEP newly classified variants, the MPNet model achieved an 85% concordance with the *FBN1* VCEP classifications. This clearly demonstrated the promise of our method and its clinical utility. Future analyses should include other disease genes with varying clinical contexts, variants of all different types and within different functional domains.

### Limitation

Several technical limitations were identified during this study. First, annotation length variability poses a significant challenge, as some variant descriptions exceed the input token limits of embedding models, resulting in the truncation of potentially relevant clinical information. This truncation can lead to the loss of critical pathogenicity indicators, particularly for complex variants with extensive functional annotations or multiple literature references. Second, despite the overall strong clustering performance, we observed instances in which variants with different pathogenicity classifications showed overlapping positions in the embedding space. This overlap occurred primarily at the boundaries between intermediate classifications, especially in suggesting that semantic similarity alone may not always capture the subtle distinctions that inform clinical decision making. Furthermore, reliance on existing variant annotations means that the approach inherits any biases or inconsistencies present in the original clinical interpretations used for training.

### Future direction

Several promising avenues exist for enhancing this approach. First, exploration of alternative embedding models, including newer transformer architectures and domain-specific biomedical models, could potentially improve clustering performance and handle annotation complexity more effectively. The integration of structured annotation formats and multimodal embedding approaches that incorporate protein structure data and functional assay results represents another significant opportunity for advancement.

The challenge of annotation length necessitates the development of specialized machine learning and deep learning frameworks designed specifically for processing variable-length clinical text. Hierarchical attention mechanisms, document-level transformers, and summarization techniques could be employed to create more robust representations that preserve critical information while maintaining computational efficiency. Additionally, the development of ensemble classifiers that combine multiple embedding approaches with traditional feature-based methods could potentially achieve superior performance compared to any single approach.

Expansion to a broader range of genes represents a critical validation step for establishing the clinical utility of this methodology. Priority should be given to genes with varying levels of clinical characterization, different functional domains, and diverse disease mechanisms to assess the universal applicability of semantic embedding approaches. Genes associated with rare diseases, where variant interpretation challenges are most acute, would provide particularly valuable validation opportunities for demonstrating the practical impact of automated classification systems in clinical genetics workflows.

## Data Availability

All data produced in the present study are available upon reasonable request to the authors.

## 5. Availability

A patent application was filed on June 10, 2025 - Predicting Genetic Variant Pathogenicity Using Vector Embeddings (U.S. Patent Application No. 63/821.249). Codes and demonstrations, however, will be released on Github.

